# Long-term patient-reported symptoms of COVID-19: an analysis of social media data

**DOI:** 10.1101/2020.07.29.20164418

**Authors:** Juan M. Banda, Gurdas Viguruji Singh, Osaid H. Alser, Daniel Prieto-Alhambra

## Abstract

As the COVID-19 virus continues to infect people across the globe, there is little understanding of the long term implications for recovered patients. There have been reports of persistent symptoms after confirmed infections on patients even after three months of initial recovery. While some of these patients have documented follow-ups on clinical records, or participate in longitudinal surveys, these datasets are usually not publicly available or standardized to perform longitudinal analyses on them. Therefore, there is a need to use additional data sources for continued follow-up and identification of latent symptoms that might be underreported in other places. In this work we present a preliminary characterization of post-COVID-19 symptoms using social media data from Twitter. We use a combination of natural language processing and clinician reviews to identify long term self-reported symptoms on a set of Twitter users.

## Introduction

As countries, such as the USA or Spain go through the perceived second wave of acute cases of COVID-19, those who suffered the condition in the preceding months are reporting concerning long-term symptoms. Acute organ injury during primo-infection includes acute kidney injury in 1 in 5 patients^1^, myocardial injury in 20%-30%, and acute respiratory failure. Long-term sequelae are therefore expected to include chronic kidney disease, heart failure and chronic obstructive pulmonary disease. As healthcare systems remain overwhelmed with the management of acute cases, little attention has been given to long-term COVID-19 sequelae.

## Methods

We mined and manually reviewed social media data from a selective set of Twitter feeds (#longcovid, #chroniccovid) to characterize patient reports of long-term COVID-19 symptoms. Using the largest publicly available COVID-19 Twitter chatter dataset^2^, we used precise hashtags (#longcovid and #chroniccovid) to select tweets relevant to discussions related to the post-COVID experiences of Twitter users. We looked at English language Twitter data from 2020-05-21 to 2020-07-10, allowing >60 days after the start of the pandemic. We discarded retweets that did not have user comments. Any tweets from accounts with unusually high tweeting activity (possible bots) or that only shared other tweets were removed. We then annotated tweets using the Social Media Mining Toolkit^3^, Spacy NER annotator and a dictionary created from the Observational Health Data Sciences and Informatics (OHDSI) vocabulary^4^, which allows the annotated terms to tie into clinical conditions and observations. We identified the final set of tweets with clinical concepts for review. Two clinicians (GS, OA) manually reviewed these tweets to identify patients with COVID-19 and their self-reported symptoms, and to attribute ICD-10 codes to them. A third clinician (DPA) reviewed all decisions and resolved disagreements. Number of symptoms per tweet and person, and frequency (%) of symptoms reported of the total were reported.

## Results

A total of 7,781 unique tweets were identified from 4,607 users. After annotation, 2,603 tweets were manually reviewed, resulting in 150 eligible tweets from 107 users. A total of 192 reports including 34 distinct ICD-10 codes were identified (Figure 1). The 10 most commonly mentioned symptoms were: malaise and fatigue (62%), dyspnea (19%), tachycardia/palpitations (13%), chest pain (13%), insomnia/sleep disorders (10%), cough (9%), headache (7%), and joint pain, fever, and unspecified pain by 6% each (Table 1).

**Table 1.** Detailed count all self-reported symptoms and the number of users presenting them.

| ICD-10 Code and Name | Users (n) |
| --- | --- |
| R53 - Malaise and fatigue | 66 |
| R06.0 - Dyspnea | 20 |
| R07.4 - Chest pain, unspecified | 14 |
| R00.0 - Tachycardia, unspecified | 14 |
| G47 - Insomnia | 11 |
| R05 - Cough | 10 |
| R51 - Headache | 7 |
| R50.9 - Fever, unspecified | 6 |
| R52 - Pain, unspecified | 6 |
| M25.5 - Pain in joint | 6 |
| M79.1 - Myalgia | 4 |
| R43.0 - Anosmia | 2 |
| R69 - Illness, unspecified | 2 |
| R41.3 - Other amnesia | 2 |
| H93.1 - Tinnitus | 2 |
| R94.1 - Abnormal results of function studies of peripheral nervous system and special senses | 2 |
| R41 - Other symptoms and signs involving cognitive functions and awareness | 1 |
| R03.0 - Elevated blood-pressure reading, without diagnosis of hypertension | 1 |
| M94 - Other disorders of cartilage | 1 |
| L29.9 - Pruritus, unspecified | 1 |
| J12.9 - Viral pneumonia, unspecified | 1 |
| H53.9 - Unspecified visual disturbance | 1 |
| D47.2 - Monoclonal gammopathy | 1 |
| R49.1 - Aphonia | 1 |
| R21 - Rash and other nonspecific skin eruption | 1 |
| R00.1 - Bradycardia, unspecified | 1 |
| J32.9 - Chronic sinusitis, unspecified | 1 |
| I26 - Pulmonary embolism | 1 |
| G90.9 - Disorder of the autonomic nervous system, unspecified | 1 |
| R68 - Other general symptoms and signs | 1 |
| R43.2 - Parageusia | 1 |
| R41.0 - Disorientation, unspecified | 1 |
| R11 - Nausea and vomiting | 1 |
| J32 - Chronic sinusitis | 1 |

**Figure 1.**
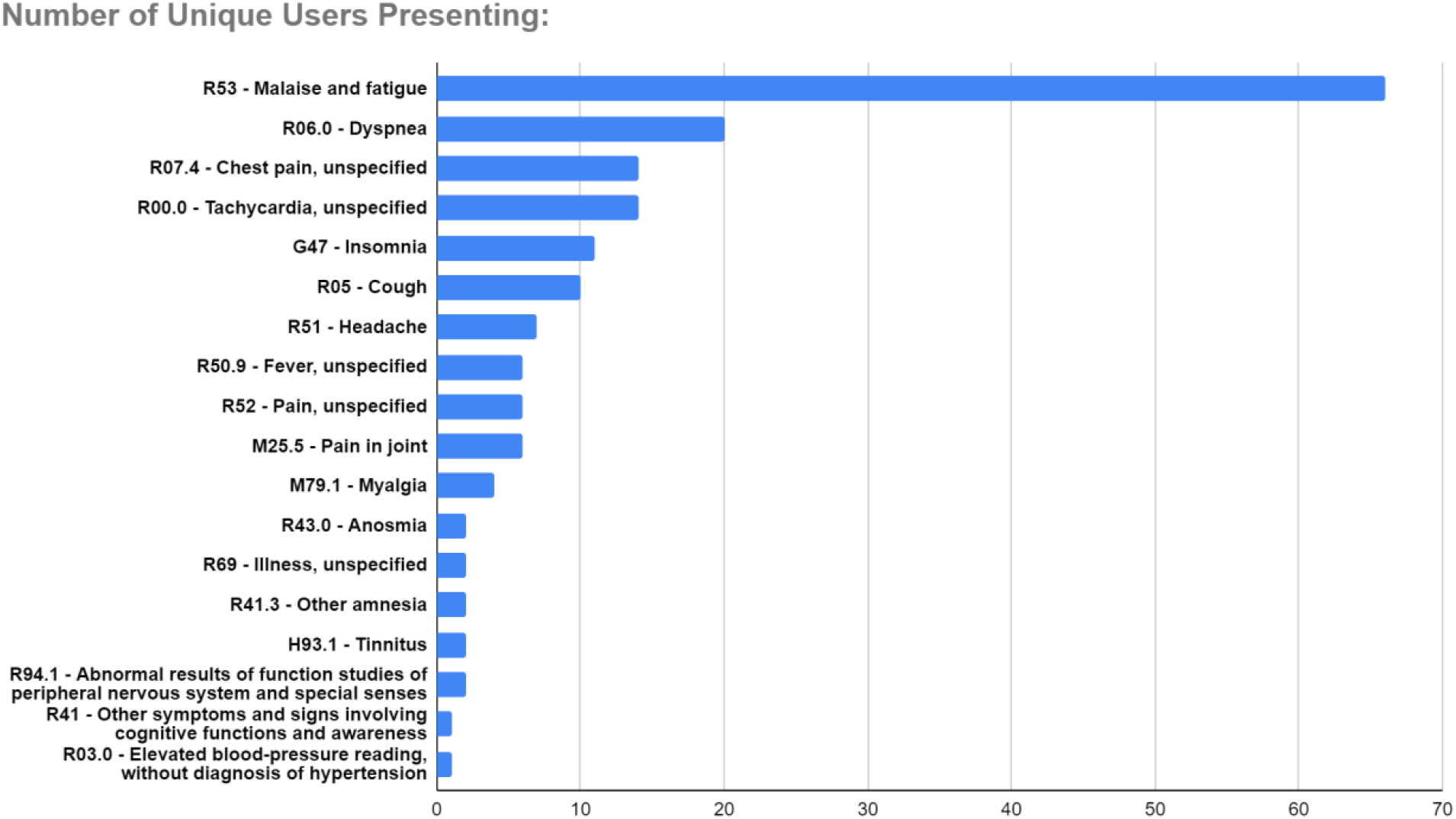
Number of users self-reporting each different symptom on their social media stream.

Less common symptoms included ear-nose-throat (tinnitus, anosmia, chronic sinusitis, parageusia, aphonia), neuro-psychological (amnesia, neuralgia/neuropathy, disautonomia, visual disturbance, cognitive impairment, and disorientation), myalgia, and skin pruritus/rash. An average of 1.79 codes were reported per person, and 1.28 codes per tweet.

## Discussion

Our analysis of patient-reported long-term COVID-19 symptoms matches clinician-collected data recently reported by Carfi *et al*^5^. Fatigue and dyspnea were the two most common symptoms in both datasets, and chest pain and cough were also reported in the top 5. Other, less commonly reported symptoms include headache, myalgia and dysgeusia/parageusia.

Patients reported additional symptoms, including palpitations and insomnia/sleep disorders potentially attributable to neuro-psychological, cardiac or respiratory sequelae. Skin pruritus and rash are increasingly recognised as COVID-19 manifestations^6^. Additional, less recognized long-term symptoms reported in our data include persistent fever, tinnitus, anosmia, and neuro-psychological distress.

We have shown that researchers can leverage social media data, specifically Twitter, to conduct long-term post-COVID studies of patient-relevant and self-reported symptoms. By leveraging the #longcovid hashtag movement, we found similar reports to those obtained from clinical adjudication^5^, demonstrating the validity of our methodological framework for future research.

Manual validation identified additional, not previously reported symptoms that warrant further research, given their importance to patients.

## Data Availability

Data will be shared upon request following the Twitter developers terms of usage.

